# Muscle Ageing and Sarcopenia Study (MASS) Lifecourse: a unique resource for understanding skeletal muscle ageing across adulthood

**DOI:** 10.64898/2026.02.18.26346577

**Authors:** Rachel Cooper, Christopher Hurst, Holly Syddall, Helen Atkinson, Jonathan G Bunn, Daniela Carpinelli, Antoneta Granic, Susan J Hillman, Emma Grace Lewis, Claire McDonald, Katie Sloan, Karen Suetterlin, Miles D Witham, Avan A Sayer

## Abstract

Advances in our understanding of the biology of skeletal muscle ageing are being made at pace, with great potential for these findings to inform the identification of novel treatments for sarcopenia. However, translation of findings from animal models to humans has been hampered by limitations of existing human muscle biopsy studies. Devised to directly address this challenge, the Muscle Ageing and Sarcopenia Study (MASS) Lifecourse is a unique resource for the study of human muscle ageing across adulthood. This deep-phenotyped observational study of 260 community-dwelling men and women aged 18 to 85 years living in North East England includes muscle biopsy samples and detailed characterisation of physical function, health status and sociodemographic and behavioural risk factors. Underpinned by broad interdisciplinary research and clinical expertise this study is catalysing cutting-edge translational research on human muscle ageing across the adult life course.

## Introduction

Skeletal muscle mass and function typically decline with age and when this process of age-related loss is accelerated, sarcopenia is diagnosed.^1, 2^ As sarcopenia is associated with myriad adverse outcomes, including loss of mobility, falls, fractures and premature mortality, it poses a major threat to healthy ageing and the maintenance of independence in later life.^3–5^ There are wide-ranging impacts of sarcopenia for individuals, their families and wider society as exemplified by an estimated annual excess cost for the care of this prevalent, but under-recognised condition, of £2.5 billion in the UK alone.^6, 7^ At the current time, resistance exercise is the only identified treatment for sarcopenia with good evidence of efficacy.^8^ As not everyone is able or willing to perform or sustain resistance exercise, novel preventive strategies and treatments for sarcopenia are needed.

In recent decades, major progress has been made towards an improved understanding of the biology of ageing.^9–11^ Consequently, hallmarks of ageing have been identified as potential treatment targets for a range of age-related conditions including sarcopenia.^12^ By studying these different hallmarks in muscle tissue our understanding of the pathophysiology of sarcopenia has advanced.^2^ However, many studies of the biology of muscle ageing have been conducted in animals and so we are at a critical juncture where well characterised studies of humans, including muscle biopsies, must be established and prioritised to ensure the timely translation of this knowledge from the bench to the bedside for patient and public benefit.^13^

Despite the invasive nature and technical challenges of undertaking muscle biopsies, muscle tissue samples have been obtained from live donors in a number of human observational studies^14, 15^ including some that include older adults and also characterise other important ageing phenotypes.^16–19^ While such research has provided useful insights, many studies have had relatively small sample sizes, an over-representation of healthy active people, narrow age ranges, a focus on a single sex and/or limited description of other potentially important characteristics, including sociodemographic and behavioural risk factors - commonly ascertained in epidemiological studies but less often in studies with a focus on biology or physiology. Limitations of existing studies present specific challenges if the desired focus is the study of muscle ageing across the life course,^20, 21^ and a life course approach is likely to be particularly illuminating given compelling evidence that both developmental and degenerative processes play a role in the aetiology of sarcopenia.^1, 2, 22, 23^

Drawing on our collective interdisciplinary expertise in academic geriatric medicine, physiology, biology, sport and exercise science and life course epidemiology we aimed to establish a deep phenotyped study that creates a unique resource for translational ageing research and supports the discovery of novel strategies to prevent and treat sarcopenia. The Muscle Ageing and Sarcopenia Study (MASS) Lifecourse was initiated in 2018 and since our protocol was published and the first 80 participants described^24^ we have increased the target sample size, extended the age range of participants to encompass the whole adult life course, and added novel measures.

As baseline data collection is now complete, we outline the full process of participant recruitment and baseline assessment, describe key characteristics of the MASS Lifecourse participants and compare the characteristics of our study participants with the general population from which they were selected. In doing so, we aim to showcase the value of the resource generated and its potential to advance understanding of human muscle ageing across the adult life course.

## Methods

### Recruitment

Between October 2018 and August 2024 (with a brief pause during 2020 due to the COVID-19 pandemic), men and women living in North East England were recruited to MASS Lifecourse. Most participants were recruited via General Practitioner (GP) practices in the North East and North Cumbria Clinical Research Network; at regular time intervals GP practices initially distributed study invitations to men and women aged 45 to 85 years who were judged to have capacity to consent to participate, with the age range later expanded to 18 to 85 years to include younger adults. To complement this main recruitment pathway, study invitations were also sent to men and women on the Newcastle National Institute for Health and Care Research (NIHR) BioResource register who had expressed an interest in taking part in clinical research. Monitoring the sex distribution of recruited participants informed ongoing purposive sampling to ensure approximately equal numbers of men and women in the final sample.

All men and women who, on receipt of a study invitation, completed and returned a reply slip indicating their willingness to participate were contacted by the study team via telephone. This screening call provided potential participants with the opportunity to ask any questions they had and enabled the study team to assess their eligibility. Exclusion criteria were kept to a minimum but, on the grounds of clinical sensitivity and safety, people who met any of the following criteria were not recruited: pregnant; diagnosed with diabetes mellitus; taking immunosuppressant, anticoagulant or antiplatelet medications (except for aspirin for primary prevention of cardiovascular disease which could be suspended); receiving palliative care; housebound; living in a residential or nursing home.

### Ethical approval and informed consent

The study was approved by the North East Newcastle & North Tyneside 1 Research Ethics Committee. Written informed consent was obtained from all participants at each phase of the study.

### Assessments

Once recruited to the study, participants’ baseline assessments were undertaken in two main stages: a home visit and a clinic visit. During the home visit (or videocall at the height of the COVID-19 pandemic), conducted by a trained member of the study team, data were collected on a broad range of sociodemographic characteristics, behavioural risk factors, self-reported health, wellbeing and cognitive function (see Table 1 for more details). In addition, participants were invited to wear a GENEActiv® Original physical activity monitor (ActivInsights Ltd., Cambridge, UK) on their dominant wrist for seven days.

**Table 1.**
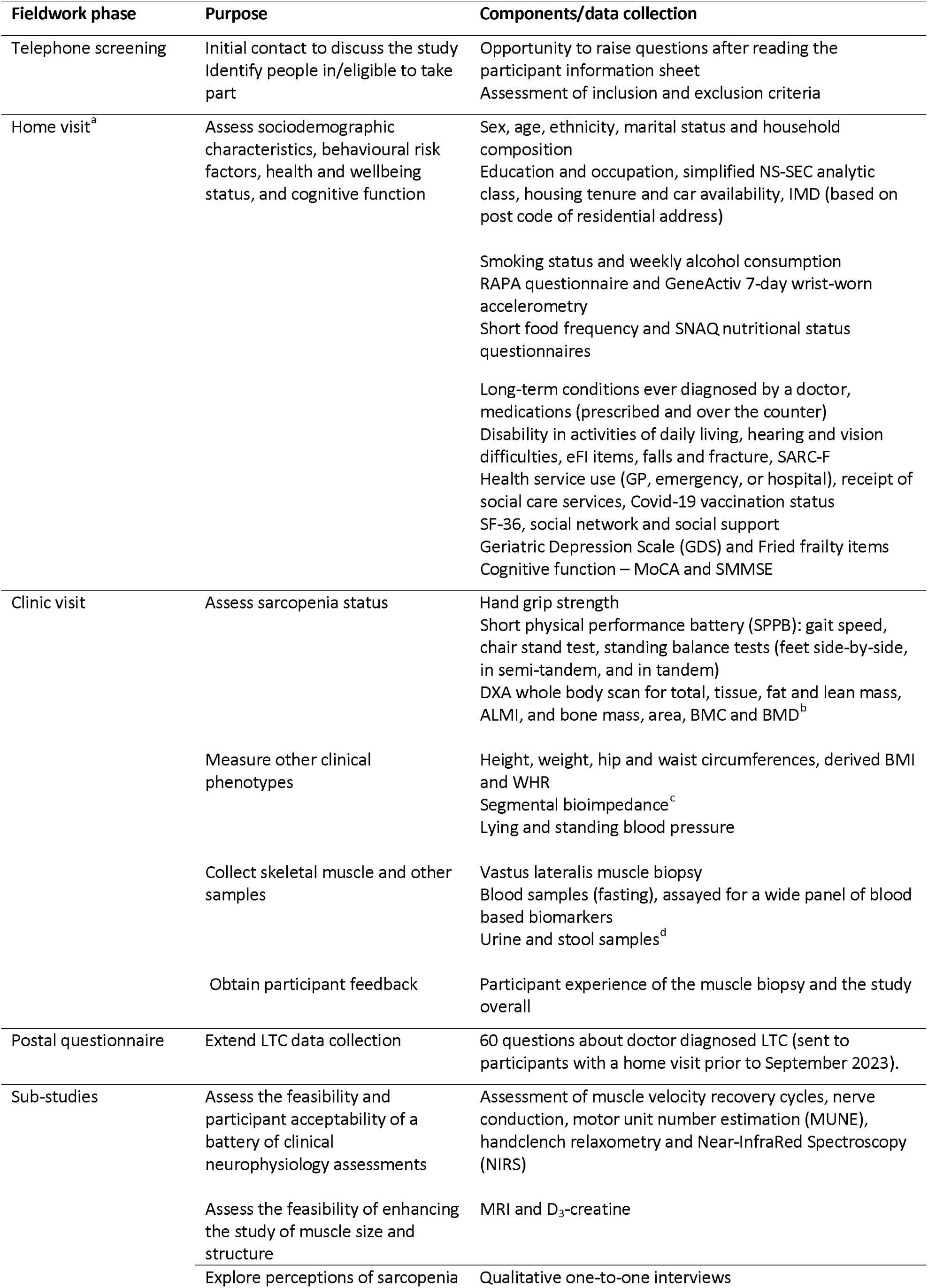
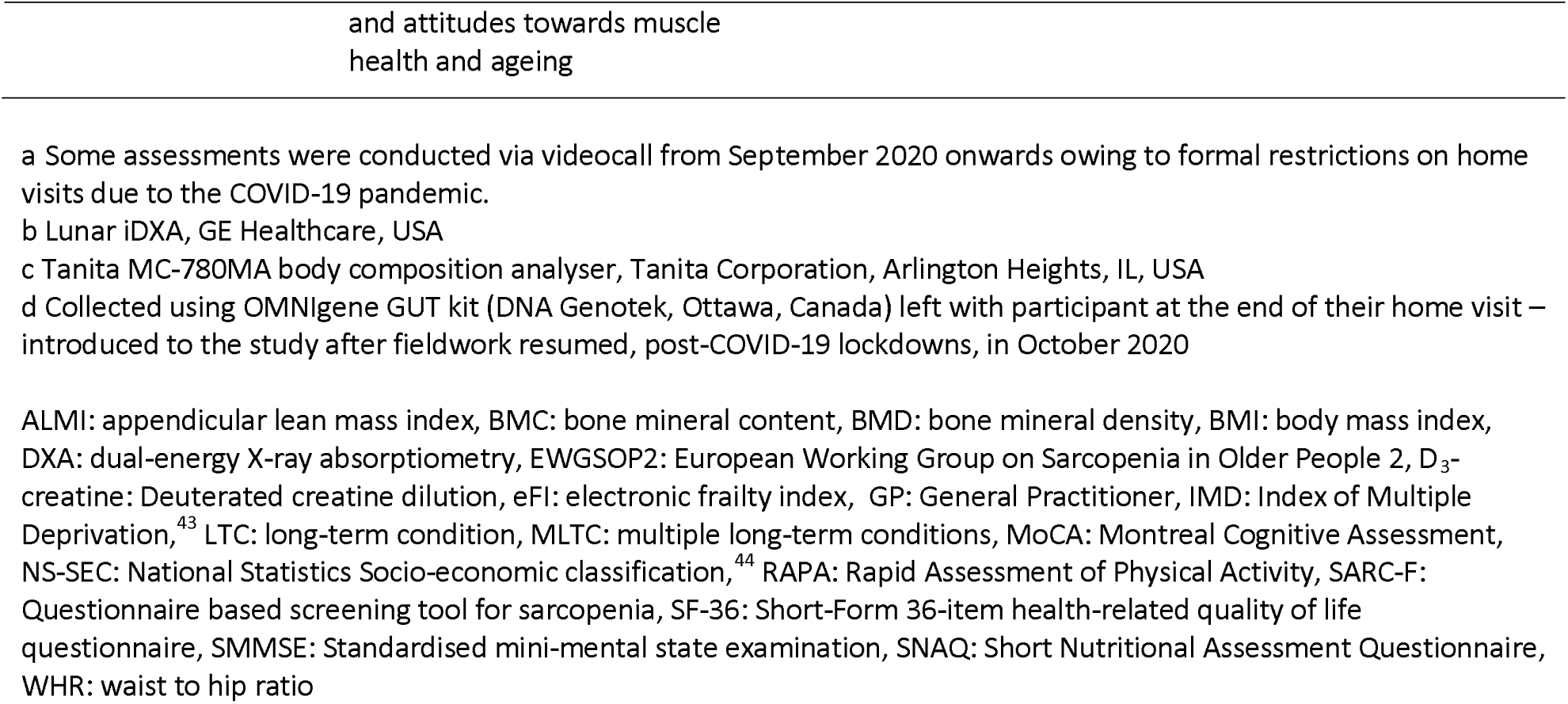
Summary of MASS Lifecourse fieldwork phases and data collected at baseline.

| Fieldwork phase | Purpose | Components/data collection |
| --- | --- | --- |
| Telephone screening | Initial contact to discuss the study<br>Identify people in/eligible to take part | Opportunity to raise questions after reading the participant information sheet<br>Assessment of inclusion and exclusion criteria |
| Home visit<sup>a</sup> | Assess sociodemographic characteristics, behavioural risk factors, health and wellbeing status, and cognitive function | Sex, age, ethnicity, marital status and household composition<br>Education and occupation, simplified NS-SEC analytic class, housing tenure and car availability, IMD (based on post code of residential address)<br>Smoking status and weekly alcohol consumption<br>RAPA questionnaire and GeneActiv 7-day wrist-worn accelerometry<br>Short food frequency and SNAQ nutritional status questionnaires<br>Long-term conditions ever diagnosed by a doctor, medications (prescribed and over the counter)<br>Disability in activities of daily living, hearing and vision difficulties, eFI items, falls and fracture, SARC-F<br>Health service use (GP, emergency, or hospital), receipt of social care services, Covid-19 vaccination status<br>SF-36, social network and social support<br>Geriatric Depression Scale (GDS) and Fried frailty items<br>Cognitive function – MoCA and SMMSE |
| Clinic visit | Assess sarcopenia status | Hand grip strength<br>Short physical performance battery (SPPB): gait speed, chair stand test, standing balance tests (feet side-by-side, in semi-tandem, and in tandem)<br>DXA whole body scan for total, tissue, fat and lean mass, ALMI, and bone mass, area, BMC and BMD<sup>b</sup> |
|  | Measure other clinical phenotypes | Height, weight, hip and waist circumferences, derived BMI and WHR<br>Segmental bioimpedance<sup>c</sup><br>Lying and standing blood pressure |
|  | Collect skeletal muscle and other samples | Vastus lateralis muscle biopsy<br>Blood samples (fasting), assayed for a wide panel of blood based biomarkers<br>Urine and stool samples<sup>d</sup> |
|  | Obtain participant feedback | Participant experience of the muscle biopsy and the study overall |
| Postal questionnaire | Extend LTC data collection | 60 questions about doctor diagnosed LTC (sent to participants with a home visit prior to September 2023). |
| Sub-studies | Assess the feasibility and participant acceptability of a battery of clinical neurophysiology assessments | Assessment of muscle velocity recovery cycles, nerve conduction, motor unit number estimation (MUNE), handclench relaxometry and Near-InfraRed Spectroscopy (NIRS) |
|  | Assess the feasibility of enhancing the study of muscle size and structure | MRI and D<sub>3</sub>-creatine |
|  | Explore perceptions of sarcopenia | Qualitative one-to-one interviews |

a Some assessments were conducted via videocall from September 2020 onwards owing to formal restrictions on home visits due to the COVID-19 pandemic.
b Lunar iDXA, GE Healthcare, USA
c Tanita MC-780MA body composition analyser, Tanita Corporation, Arlington Heights, IL, USA
d Collected using OMNIgene GUT kit (DNA Genotek, Ottawa, Canada) left with participant at the end of their home visit – introduced to the study after fieldwork resumed, post-COVID-19 lockdowns, in October 2020
ALMI: appendicular lean mass index, BMC: bone mineral content, BMD: bone mineral density, BMI: body mass index, DXA: dual-energy X-ray absorptiometry, EWGSOP2: European Working Group on Sarcopenia in Older People 2, D<sub>3</sub>-creatine: Deuterated creatine dilution, eFI: electronic frailty index, GP: General Practitioner, IMD: Index of Multiple Deprivation,<sup>43</sup> LTC: long-term condition, MLTC: multiple long-term conditions, MoCA: Montreal Cognitive Assessment, NS-SEC: National Statistics Socio-economic classification,<sup>44</sup> RAPA: Rapid Assessment of Physical Activity, SARC-F: Questionnaire based screening tool for sarcopenia, SF-36: Short-Form 36-item health-related quality of life questionnaire, SMMSE: Standardised mini-mental state examination, SNAQ: Short Nutritional Assessment Questionnaire, WHR: waist to hip ratio

Participants were then invited to undertake a comprehensive clinical assessment at the Clinical Ageing Research Unit (CARU) within the NIHR Newcastle Clinical Research Facility. During this research nurse-led visit, the following assessments were conducted using standardised protocols^25, 26^: grip strength, physical performance, body composition, and blood pressure. In addition, skeletal muscle biopsy, fasting blood, urine and stool samples were collected.

Biopsy of the vastus lateralis muscle (usually right leg) was conducted by a trained clinician using a Weil-Blakesley conchotome.^27^ Vastus lateralis was chosen because it is an accessible site which avoids the main neurovascular structures in the thigh,^27^ and we have previously demonstrated its feasibility and acceptability.^28, 29^ Three muscle samples were collected, with one placed immediately into RNAlater solution (RNAlater™ stabilization solution, Thermofisher Scientific). The other two samples were placed on sterile gauze for transportation prior to processing: one sample orientated and mounted for histology, and then both samples flash frozen in isopentane cooled in liquid nitrogen prior to storage at − 80◦C.^30^ The participant was observed for approximately two hours after the biopsy and followed up by telephone twice, 24-48 hours and one week later.

At the end of this clinic visit, participants were asked to provide feedback on their experience, and whether they agreed to being contacted again regarding future research opportunities.

### Additional assessments to enhance the resource

During later stages of the baseline assessment period, a standardised approach to the definition and operationalisation of multiple long-term conditions (MLTC) was developed by members of the study team.^31^ This recommended including 60 specific LTC when defining MLTC, only 25 of which had been assessed in home visits up to August 2023. Postal questionnaires were therefore prepared and sent to study participants whose home visit took place before August 2023 to ascertain information on all 60 LTC. Study participants who were recruited from August 2023 onwards were asked about this extended set of 60 LTC during their home visit.

Reflecting its potential as a platform for novel sub-studies of muscle ageing, during an early phase of baseline assessment a standalone study of Phase Contrast Motor Unit Magnetic Resonance Imaging (PC-MUMRI) in the calf was undertaken in 61 MASS Lifecourse participants by collaborators.^32^ During a later phase (March-July 2024), a sub-set of participants were invited to contribute to an additional clinical assessment that took place before the main clinic assessment and muscle biopsy. This included assessing the feasibility of incorporating established and novel clinical neurophysiology measures and additional assessments of muscle size and structure (using magnetic resonance imaging (MRI) and Deuterated creatine (D_3_-creatine) dilution). A qualitative study has also been initiated, which involves in depth one-to-one interviews with a small sample of study participants to explore perceptions of sarcopenia and attitudes towards muscle health and ageing.

### Patient and public involvement

Patients and the public who have shared their lived experience of muscle ageing^2^ with our study team during clinical interactions and public engagement events provided the motivation for this study. Once recruited, engagement with study participants has been maintained via an annual newsletter and an in-person event.^33^ This flagship event gave the study team the opportunity to thank participants for their contributions, disseminate early findings and seek feedback on the acceptability of plans for follow-up assessments. Participants and public contributors to an advisory panel remain involved in plans for future phases of the study.

### Statistical analyses

After conducting quality control checks of all data, study participant characteristics have been described by sex, using medians and inter-quartile ranges (25th to 75th percentiles) for continuously distributed variables, and frequencies and percentage distributions for binary and categorical variables, using the maximum available sample size for each variable. To assess its construct validity, mean maximum grip strength was described by sex and decades of age.

To allow us to assess how MASS Lifecourse participants compare with the wider populations from which they were selected at the level of local authority of residence (Northumberland, North Tyneside, or Newcastle), region (North East England), and country (England), key characteristics in MASS Lifecourse and these other sources were described for men and women combined.^34–37^

Statistical analyses were conducted using StataNow, release 18.5.^38^

## Results

Of 3244 study invitations distributed, a total of 715 responses (22%) were received (see Figure 1). Of these, 396 people (55%) expressed an interest in participating and underwent telephone screening. After exclusion of 45 people for medical reasons, a total of 351 (89%) people were identified as eligible and invited to participate in a home visit, of whom 260 (74%) were recruited and underwent the assessments outlined in Table 1.

**Figure 1.**
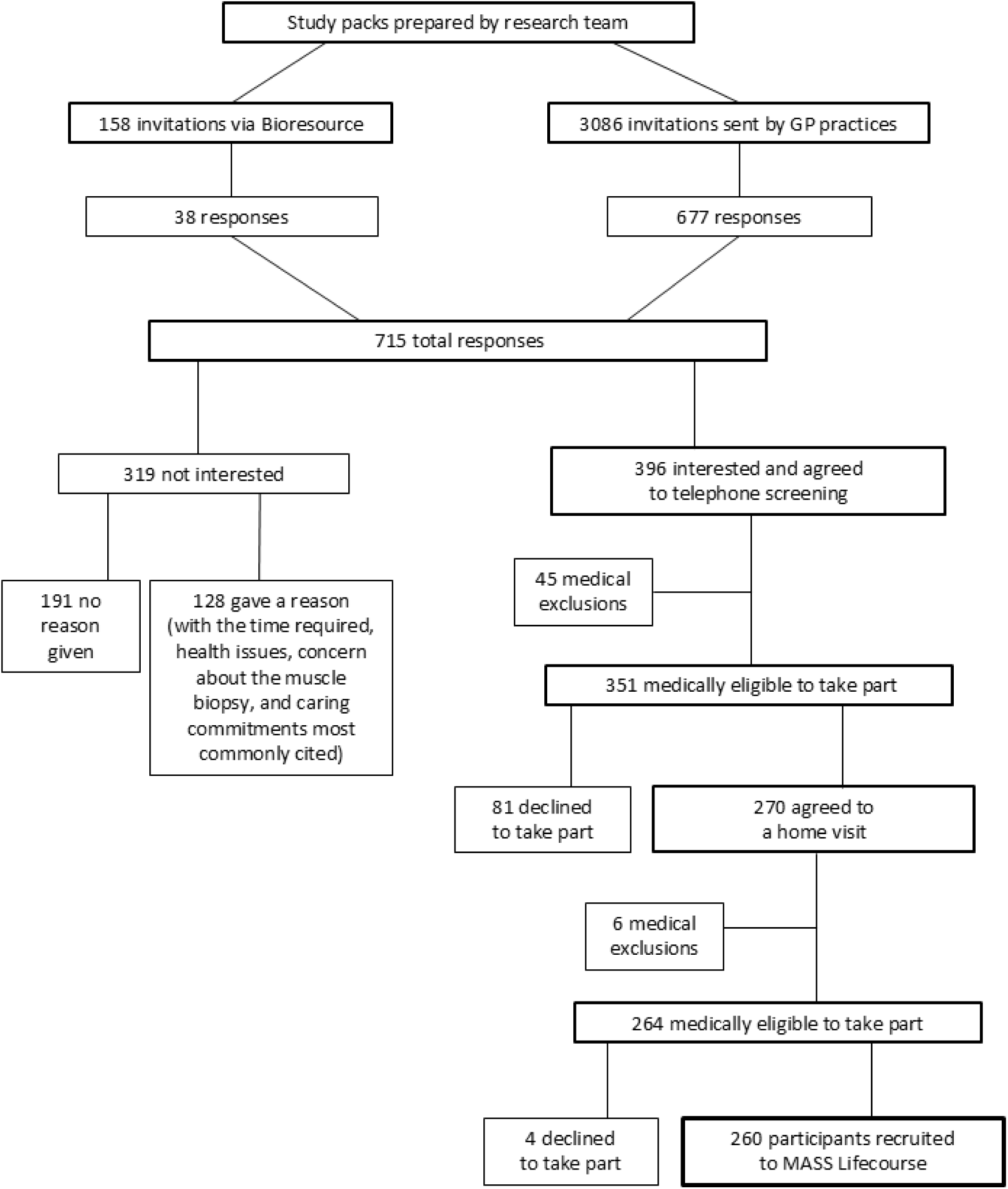
Recruitment of participants to MASS Lifecourse.

The 260 participants recruited (129 men, 131 women) were aged 18 to 85 years but by design the large majority were aged 50 years or older (see Table 2). Nearly all participants were of white ethnicity, approximately half had remained in education post-16 years of age, and 26% were living in the least deprived neighbourhoods (as indicated by being in the top tenth of the index of multiple deprivation). Few participants reported being a current smoker, although approximately a third were ex-smokers. Over two-thirds of participants rated their general health as excellent or very good (see Table 3) despite self-reported doctor diagnosed long-term conditions (LTC) being common: 26% of men and 28% of women reported a single LTC, and 54% of men and 58% of women reported two or more long-term conditions (referred to as multiple long-term conditions (MLTC)). In addition to this 69% of participants were taking at least one prescription medication, with 7% taking five or more.

**Table 2.**
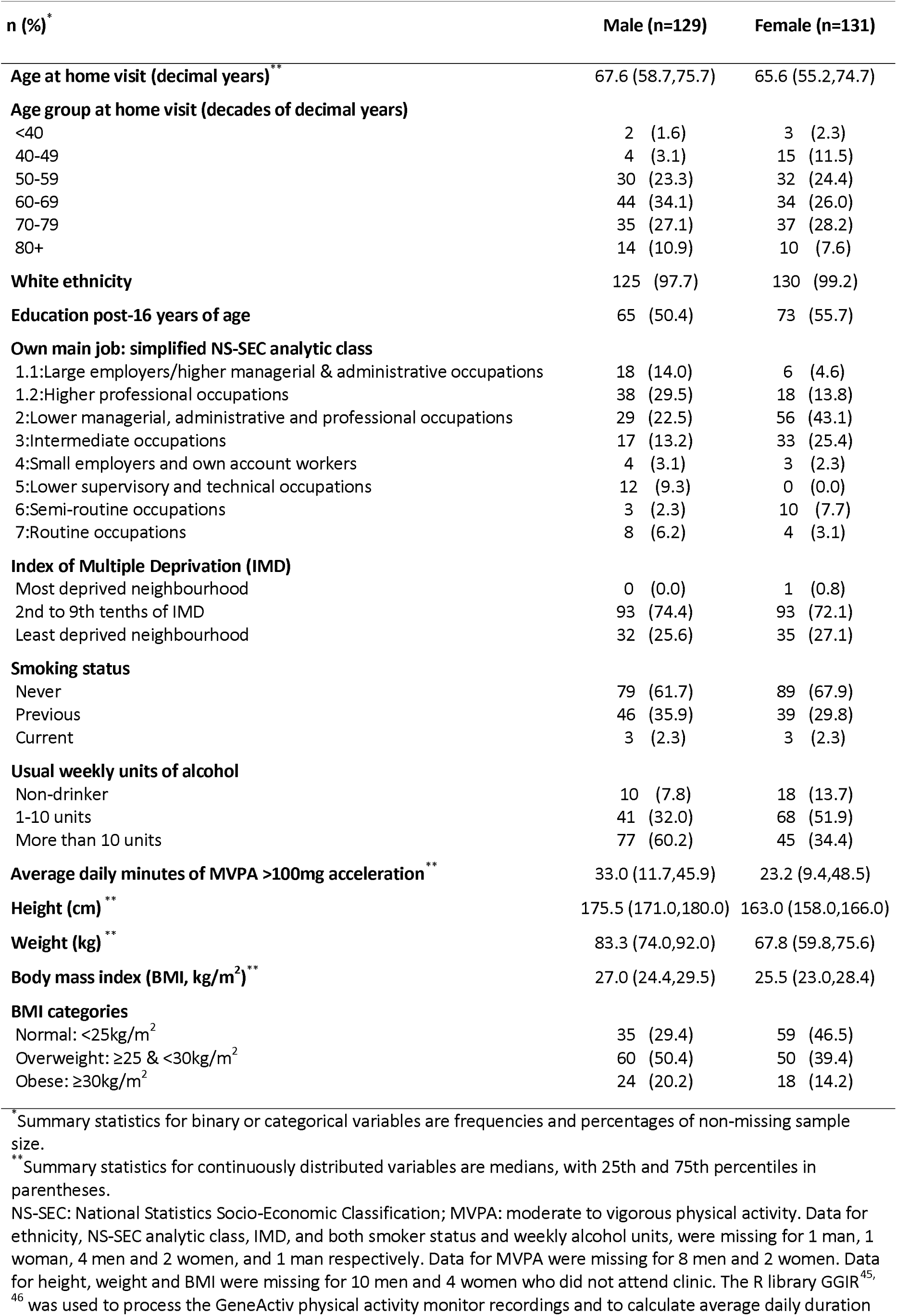
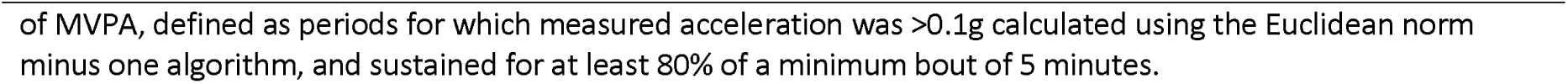
Characteristics of MASS Lifecourse participants at baseline by sex.

| n (%)<sup>*</sup> | Male (n=129) | Female (n=131) |
| --- | --- | --- |
| **Age at home visit (decimal years)<sup>\*\*</sup>** | 67.6 (58.7,75.7) | 65.6 (55.2,74.7) |
| **Age group at home visit (decades of decimal years)** |  |  |
| <40 | 2 (1.6) | 3 (2.3) |
| 40-49 | 4 (3.1) | 15 (11.5) |
| 50-59 | 30 (23.3) | 32 (24.4) |
| 60-69 | 44 (34.1) | 34 (26.0) |
| 70-79 | 35 (27.1) | 37 (28.2) |
| 80+ | 14 (10.9) | 10 (7.6) |
| **White ethnicity** | 125 (97.7) | 130 (99.2) |
| **Education post-16 years of age** | 65 (50.4) | 73 (55.7) |
| **Own main job: simplified NS-SEC analytic class** |  |  |
| 1.1: Large employers/higher managerial & administrative occupations | 18 (14.0) | 6 (4.6) |
| 1.2: Higher professional occupations | 38 (29.5) | 18 (13.8) |
| 2: Lower managerial, administrative and professional occupations | 29 (22.5) | 56 (43.1) |
| 3: Intermediate occupations | 17 (13.2) | 33 (25.4) |
| 4: Small employers and own account workers | 4 (3.1) | 3 (2.3) |
| 5: Lower supervisory and technical occupations | 12 (9.3) | 0 (0.0) |
| 6: Semi-routine occupations | 3 (2.3) | 10 (7.7) |
| 7: Routine occupations | 8 (6.2) | 4 (3.1) |
| **Index of Multiple Deprivation (IMD)** |  |  |
| Most deprived neighbourhood | 0 (0.0) | 1 (0.8) |
| 2nd to 9th tenths of IMD | 93 (74.4) | 93 (72.1) |
| Least deprived neighbourhood | 32 (25.6) | 35 (27.1) |
| **Smoking status** |  |  |
| Never | 79 (61.7) | 89 (67.9) |
| Previous | 46 (35.9) | 39 (29.8) |
| Current | 3 (2.3) | 3 (2.3) |
| **Usual weekly units of alcohol** |  |  |
| Non-drinker | 10 (7.8) | 18 (13.7) |
| 1-10 units | 41 (32.0) | 68 (51.9) |
| More than 10 units | 77 (60.2) | 45 (34.4) |
| **Average daily minutes of MVPA >100mg acceleration<sup>\*\*</sup>** | 33.0 (11.7,45.9) | 23.2 (9.4,48.5) |
| **Height (cm)<sup>\*\*</sup>** | 175.5 (171.0,180.0) | 163.0 (158.0,166.0) |
| **Weight (kg)<sup>\*\*</sup>** | 83.3 (74.0,92.0) | 67.8 (59.8,75.6) |
| **Body mass index (BMI, kg/m<sup>2</sup>)<sup>\*\*</sup>** | 27.0 (24.4,29.5) | 25.5 (23.0,28.4) |
| **BMI categories** |  |  |
| Normal: <25kg/m<sup>2</sup> | 35 (29.4) | 59 (46.5) |
| Overweight: ≥25 & <30kg/m<sup>2</sup> | 60 (50.4) | 50 (39.4) |
| Obese: ≥30kg/m<sup>2</sup> | 24 (20.2) | 18 (14.2) |

<sup>\*</sup> Summary statistics for binary or categorical variables are frequencies and percentages of non-missing sample size.
<sup>\*\*</sup> Summary statistics for continuously distributed variables are medians, with 25th and 75th percentiles in parentheses.
NS-SEC: National Statistics Socio-Economic Classification; MVPA: moderate to vigorous physical activity. Data for ethnicity, NS-SEC analytic class, IMD, and both smoker status and weekly alcohol units, were missing for 1 man, 1 woman, 4 men and 2 women, and 1 man respectively. Data for MVPA were missing for 8 men and 2 women. Data for height, weight and BMI were missing for 10 men and 4 women who did not attend clinic. The R library GGIR<sup>45</sup>,
<sup>46</sup> was used to process the GeneActiv physical activity monitor recordings and to calculate average daily duration
of MVPA, defined as periods for which measured acceleration was $>0.1g$ calculated using the Euclidean norm minus one algorithm, and sustained for at least 80% of a minimum bout of 5 minutes.

**Table 3.** Health status of MASS Lifecourse participants at baseline by sex.

| **n (%)<sup>\*</sup>** | **Male (n=129)** | **Female (n=131)** |
| --- | --- | --- |
| **Number of long-term conditions (LTC) <sup>\*\*</sup>** |  |  |
| None | 26 (20.2) | 19 (14.5) |
| One | 33 (25.6) | 36 (27.5) |
| Two or more (MLTC) | 70 (54.3) | 76 (58.0) |
| **Number of prescribed medications** |  |  |
| None | 42 (32.6) | 39 (29.8) |
| 1-4 | 78 (60.5) | 82 (62.6) |
| 5 or more (polypharmacy) | 9 (7.0) | 10 (7.6) |
| **SF-36 Self-rated general health** |  |  |
| Excellent | 22 (17.1) | 32 (24.4) |
| Very good | 64 (49.6) | 62 (47.3) |
| Good | 40 (31.0) | 30 (22.9) |
| Fair | 2 (1.6) | 6 (4.6) |
| Poor | 1 (0.8) | 1 (0.8) |
| **Number of falls in the past year** |  |  |
| None | 117 (90.7) | 103 (78.6) |
| One | 9 (7.0) | 21 (16.0) |
| 2 or more | 3 (2.3) | 7 (5.3) |
| **Geriatric Depression Scale (GDS)** |  |  |
| Score (range 0-15) | 0.0 (0.0,1.0) | 0.0 (0.0,1.0) |
| Possible depression (score ≥5) | 2 (1.6) | 6 (4.6) |
| **Montreal Cognitive Assessment (MoCA)** |  |  |
| Total Score (range 0-30) | 27.0 (25.0,28.0) | 27.0 (26.0,29.0) |
| Poor cognition (score <26) | 44 (34.1) | 32 (24.4) |
| **Health service use** |  |  |
| GP contact (in last four weeks) | 28 (21.9) | 19 (14.5) |
| Emergency care (ambulance or A&E in last 3 months) | 6 (4.7) | 6 (4.6) |
| Hospital outpatient (in last 3 months) | 24 (18.8) | 22 (16.8) |
| Hospital day patient (in past year) | 21 (16.4) | 19 (14.5) |
| Hospital inpatient (in past year) | 2 (1.6) | 4 (3.1) |
| **SARC-F questionnaire score** |  |  |
| 0 | 113 (87.6) | 88 (67.2) |
| 1 | 13 (10.1) | 36 (27.5) |
| 2 | 0 (0.0) | 3 (2.3) |
| 3 | 2 (1.6) | 1 (0.8) |
| 4+ | 1 (0.8) | 3 (2.3) |

<sup>\*</sup> Summary statistics are frequencies and percentages of non-missing sample size for binary or categorical variables, and are medians (with 25th and 75th percentiles in parentheses) for continuously distributed variables.
<sup>\*\*</sup> Based on the list of 60 specific LTC recommended by Cooper et al for the standardised operationalisation of multiple long-term conditions (MLTC).<sup>31</sup>
LTC: long-term condition; MLTC: multiple long-term conditions; SF-36: Short-Form 36-item health-related quality of life questionnaire; GDS: Geriatric Depression Scale; MoCA: Montreal Cognitive Assessment; GP: General Practitioner; A&E: Accident and Emergency department; SARC-F: Strength, assistance with walking, rising from a chair, climbing stairs, and falls. 1 man had missing data for the GDS and health service use variables.

Of the 260 participants who completed a home visit, 246 (95%) (119 men, 127 women) also completed a clinic visit, during which a muscle biopsy was undertaken for 239 (116 men, 123 women). Descriptive analyses showed that women had lower average grip strength than men (see Table 4) and in both sexes average grip strength declined progressively with increasing age, confirming construct validity (see Figure 2). Appendicular lean mass index (ALMI) was also lower in women than men, but average walking speed and time taken to complete five chair stands were similar (see Table 4). When European Working Group on Sarcopenia in Older People (second consensus, EWGSOP2) cut-points^39^ were applied to these continuous measures a higher proportion of women than men were classified as having low muscle strength and low lean mass (see Table 4). Prevalence of probable and confirmed sarcopenia, defined using EWGSOP2 criteria, varied depending on whether or not low muscle strength was classified according to low grip strength only, or low grip strength and/or slow chair stand speed. When using the grip strength criterion only, 3.4% of men and 6.3% of women were classified as having probable sarcopenia and no men and 1.6% of women had confirmed sarcopenia. When using grip strength or chair stands criteria, estimates increased to 21.8% of men and 16.7% of women with probable sarcopenia and no men and 6.3% of women with confirmed sarcopenia.

**Figure 2.**
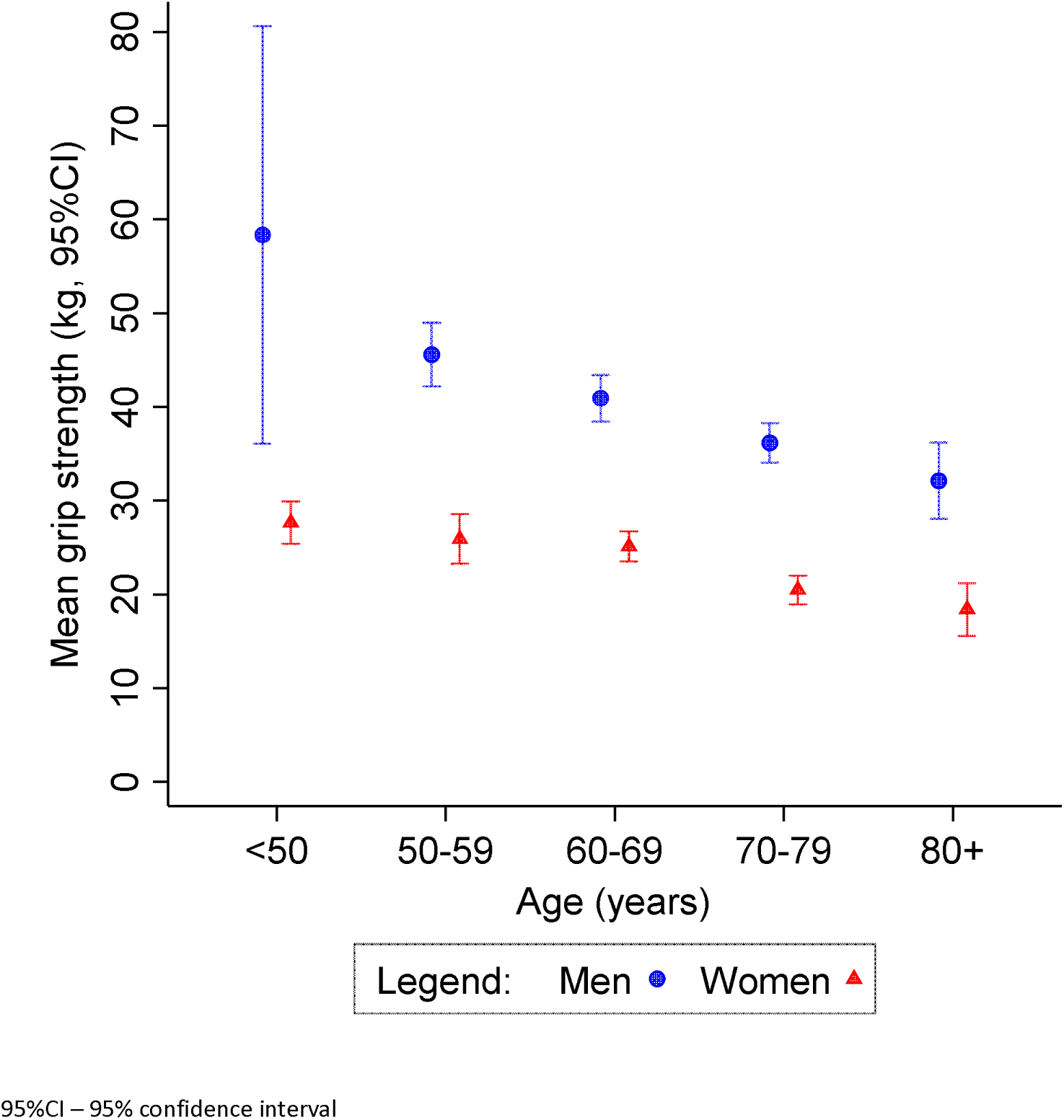
Mean grip strength by sex and age in MASS Lifecourse participants (119 men and 126 women)

**Table 4.** Muscle ageing characteristics of MASS Lifecourse participants at baseline by sex.

| n (%)<sup>*</sup> | Male (n=119) | Female (n=127) |
| --- | --- | --- |
| **Grip strength** |  |  |
| Maximum (kg)<sup>**</sup> | 40.0 (32.0,46.0) | 24.0 (20.0,28.0) |
| EWGSOP2 low strength (M<27kg; F<16kg) | 4 (3.4) | 10 (7.9) |
| **Chair stands** |  |  |
| Time to complete five stands (s)<sup>**</sup> | 12.0 (9.6,14.4) | 11.4 (9.5,14.2) |
| EWGSOP2 slow rate (M&F >15s for five stands) | 22 (18.5) | 25 (19.8) |
| **Appendicular lean mass** |  |  |
| Appendicular lean mass index (ALMI, kg/m<sup>2</sup>)<sup>**</sup> | 8.2 (7.6,8.8) | 6.3 (5.9,6.9) |
| EWGSOP2 low ALMI (M<7.0kg/m<sup>2</sup>; F<5.5kg/m<sup>2</sup>) | 8 (6.7) | 17 (13.4) |
| **Gait speed** |  |  |
| Speed over 4m (m/s)<sup>**</sup> | 1.1 (1.0,1.3) | 1.2 (1.0,1.3) |
| EWGSOP2 slow gait speed (M&F ≤0.8m/s) | 9 (7.6) | 9 (7.1) |
| **Short physical performance battery (SPPB)<sup>\*\*\*</sup>** |  |  |
| Score (range 0-12)<sup>**</sup> | 11 (10,12) | 11 (10,12) |
| EWGSOP2 low SPPB score (M&F ≤8) | 5 (4.2) | 10 (7.9) |
| **Sarcopenia** |  |  |
| *EWGSOP2 - grip strength criterion only* |  |  |
| None | 115 (96.6) | 116 (92.1) |
| Probable | 4 (3.4) | 8 (6.3) |
| Confirmed | 0 (0.0) | 2 (1.6) |
| *EWGSOP2 - grip strength or chair stands criteria* |  |  |
| None | 93 (78.2) | 97 (77.0) |
| Probable | 26 (21.8) | 21 (16.7) |
| Confirmed | 0 (0.0) | 8 (6.3) |

<sup>\*</sup>Summary statistics for binary or categorical variables are frequencies and percentages of non-missing sample size.
<sup>\*\*</sup>Summary statistics for continuously distributed variables are medians, with 25th and 75th percentiles in parentheses.
<sup>\*\*\*</sup>Short physical performance battery (SPPB) score derived from performance on tests of gait speed, chair stands, and standing balance tests (feet side-by-side, in semi-tandem, and in tandem) as per Guralnik et al<sup>47</sup>
EWGSOP2 – European Working Group on Sarcopenia in Older People 2; M – male; F – female; m/s – metres per second; s – seconds; m – metres; SPPB – short physical performance battery. All the measures described in this table were assessed at the MASS Lifecourse clinic assessment which was attended by 119 men and 127 women. One man had missing data for semi-tandem and tandem standing balance times and consequently had missing data for SPPB score. One woman had missing data for all but ALMI and consequently had missing data for SPPB score and sarcopenia.

In their study feedback (Supplementary Material, Table 1), 63% of men and 50% of women reported that the biopsy was better than they expected and only 2% reported that it was worse. Reassuringly, given the value of longitudinal assessment and plans for follow-up, 97% of men and 98% of women said that they would be willing to have another muscle biopsy for research purposes in the future.

The study’s recruitment strategy yielded a sample of participants which was equally split between men and women, but in which older people were over-represented by design. The majority of MASS Lifecourse participants self-identified as being white British, which is comparable with the populations of the Northumberland and North Tyneside local authority areas from which 91% of the study participants were recruited (see Table 5). A lower proportion of study participants were current smokers than comparator populations, but the proportions overweight or obese were similar. The least deprived tenth of the index of multiple deprivation was over-represented among MASS Lifecourse participants, but despite this, the leading causes of morbidity were commensurate with those in comparator populations.

**Table 5.**
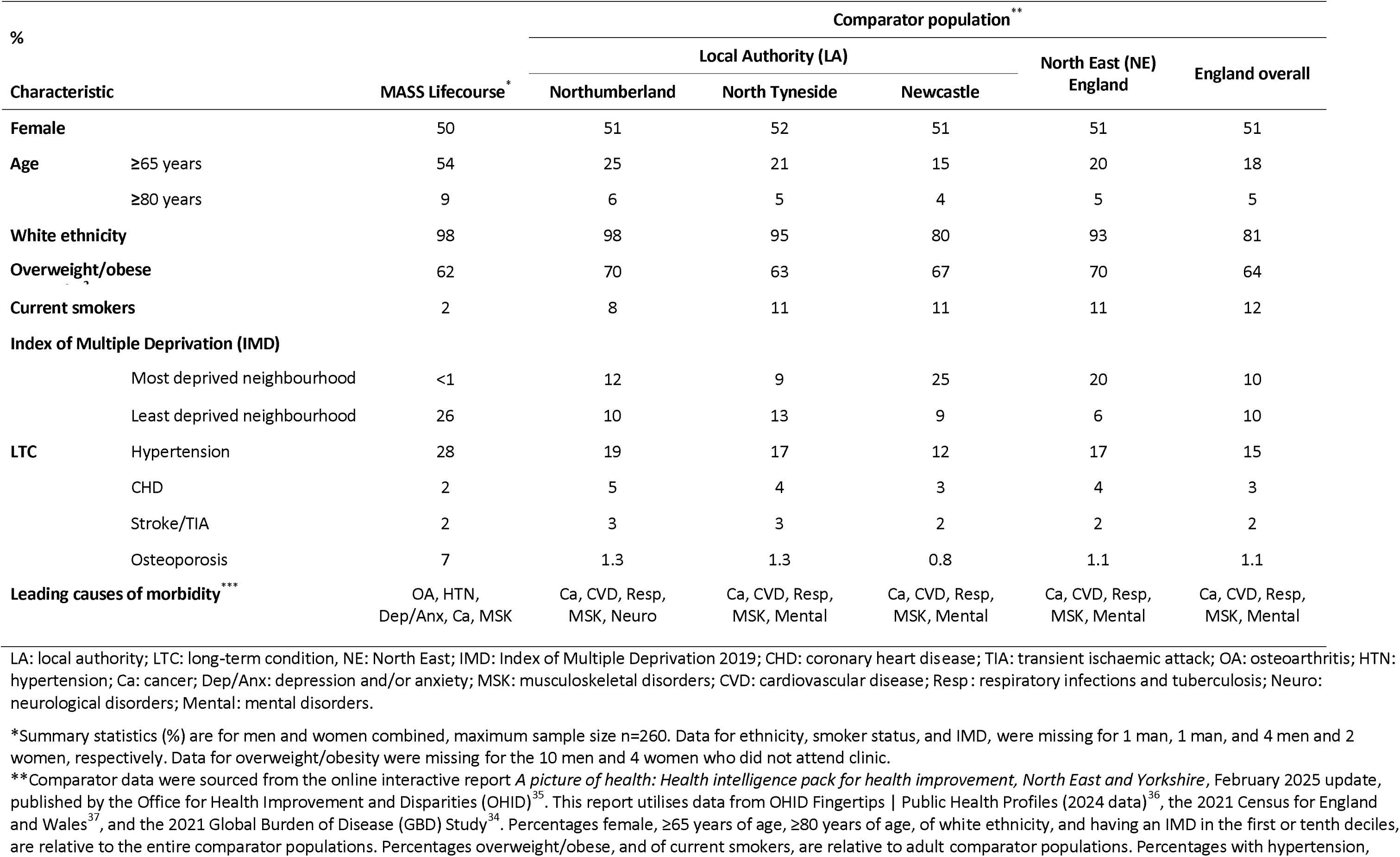
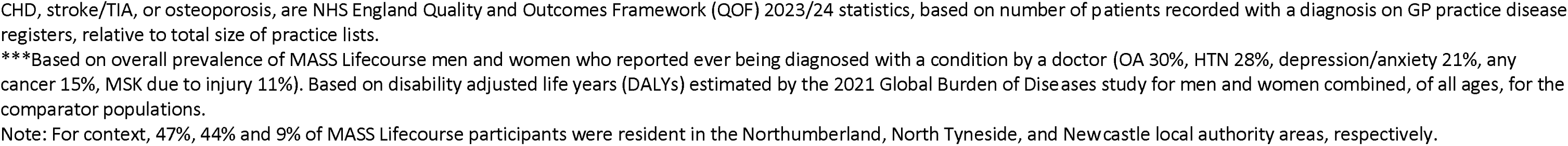
Characteristics of MASS Lifecourse participants and comparator populations at local authority, regional, and country level.

| % |  | Comparator population ** |  |  |  |  |
| --- | --- | --- | --- | --- | --- | --- |
|  |  | Local Authority (LA) |  |  | North East (NE)<br>England | England overall |
| Characteristic | MASS Lifecourse * | Northumberland | North Tyneside | Newcastle |  |  |
| **Female** | 50 | 51 | 52 | 51 | 51 | 51 |
| **Age** |  |  |  |  |  |  |
| ≥65 years | 54 | 25 | 21 | 15 | 20 | 18 |
| ≥80 years | 9 | 6 | 5 | 4 | 5 | 5 |
| **White ethnicity** | 98 | 98 | 95 | 80 | 93 | 81 |
| **Overweight/obese** | 62 | 70 | 63 | 67 | 70 | 64 |
| **Current smokers** | 2 | 8 | 11 | 11 | 11 | 12 |
| **Index of Multiple Deprivation (IMD)** |  |  |  |  |  |  |
| Most deprived neighbourhood | <1 | 12 | 9 | 25 | 20 | 10 |
| Least deprived neighbourhood | 26 | 10 | 13 | 9 | 6 | 10 |
| **LTC** |  |  |  |  |  |  |
| Hypertension | 28 | 19 | 17 | 12 | 17 | 15 |
| CHD | 2 | 5 | 4 | 3 | 4 | 3 |
| Stroke/TIA | 2 | 3 | 3 | 2 | 2 | 2 |
| Osteoporosis | 7 | 1.3 | 1.3 | 0.8 | 1.1 | 1.1 |
| **Leading causes of morbidity \*\*\*** | OA, HTN,<br>Dep/Anx, Ca, MSK | Ca, CVD, Resp,<br>MSK, Neuro | Ca, CVD, Resp,<br>MSK, Mental | Ca, CVD, Resp,<br>MSK, Mental | Ca, CVD, Resp,<br>MSK, Mental | Ca, CVD, Resp,<br>MSK, Mental |

LA: local authority; LTC: long-term condition, NE: North East; IMD: Index of Multiple Deprivation 2019; CHD: coronary heart disease; TIA: transient ischaemic attack; OA: osteoarthritis; HTN: hypertension; Ca: cancer; Dep/Anx: depression and/or anxiety; MSK: musculoskeletal disorders; CVD: cardiovascular disease; Resp: respiratory infections and tuberculosis; Neuro: neurological disorders; Mental: mental disorders.
\*Summary statistics (%) are for men and women combined, maximum sample size n=260. Data for ethnicity, smoker status, and IMD, were missing for 1 man, 1 man, and 4 men and 2 women, respectively. Data for overweight/obesity were missing for the 10 men and 4 women who did not attend clinic.
\*\*Comparator data were sourced from the online interactive report *A picture of health: Health intelligence pack for health improvement, North East and Yorkshire*, February 2025 update, published by the Office for Health Improvement and Disparities (OHID)<sup>35</sup>. This report utilises data from OHID Fingertips | Public Health Profiles (2024 data)<sup>36</sup>, the 2021 Census for England and Wales<sup>37</sup>, and the 2021 Global Burden of Disease (GBD) Study<sup>34</sup>. Percentages female, ≥65 years of age, ≥80 years of age, of white ethnicity, and having an IMD in the first or tenth deciles, are relative to the entire comparator populations. Percentages overweight/obese, and of current smokers, are relative to adult comparator populations. Percentages with hypertension,
CHD, stroke/TIA, or osteoporosis, are NHS England Quality and Outcomes Framework (QOF) 2023/24 statistics, based on number of patients recorded with a diagnosis on GP practice disease registers, relative to total size of practice lists.
\*\*\*Based on overall prevalence of MASS Lifecourse men and women who reported ever being diagnosed with a condition by a doctor (OA 30%, HTN 28%, depression/anxiety 21%, any cancer 15%, MSK due to injury 11%). Based on disability adjusted life years (DALYs) estimated by the 2021 Global Burden of Diseases study for men and women combined, of all ages, for the comparator populations.
Note: For context, 47%, 44% and 9% of MASS Lifecourse participants were resident in the Northumberland, North Tyneside, and Newcastle local authority areas, respectively.

## Discussion

Advances in our understanding of the biology of skeletal muscle ageing are being made at pace,^2, 9, 21^ but our ability to translate findings from animal models to humans has been hampered by the limitations of many human muscle biopsy studies to date. MASS Lifecourse, a deep phenotyped cohort of 260 community-dwelling adults aged 18 to 85 years living in North East England, has been established to directly address these limitations and is a unique resource for the study of muscle ageing across adulthood. Drawing on broad interdisciplinary research and clinical expertise, MASS Lifecourse provides novel opportunities for the scientific community to deliver cutting edge translational research that will advance our understanding of human muscle ageing across the adult life course and generate robust evidence to inform the identification of novel preventive strategies and treatments for sarcopenia. It explicitly addresses a recognised barrier to progress in the field of muscle ageing research, that ‘is the limited access to healthy human tissue across the lifespan’ which has been described by Kedlian et al (p736) as ‘the bottleneck for human studies’.^21^

An early example of the potential of this study to catalyse translational research is the use of MASS Lifecourse samples to interrogate the evidence of cellular senescence identified in ageing skeletal mouse muscle^40^ in human muscle tissue.^41^ Now that baseline assessments are complete, further research to accelerate the translation of important discoveries in ageing biology into studies of well characterised humans can be advanced via collaborations with a growing network of interdisciplinary researchers and industry partners.

Few human observational studies, with muscle tissue sample collection, have the breadth and depth of data on such a wide range of other relevant characteristics across the full adult age range as MASS Lifecourse. This study therefore complements and adds value to the small portfolio of other studies globally^16–19^ that together present exciting opportunities to catalyse translational research on ageing muscle across the life course, identify novel treatment targets and deliver benefits for patients and the public.

Recording the number of community-dwelling adults who needed to be contacted to achieve a recruitment target of 260 willing to undergo time consuming and invasive assessments, including a muscle biopsy, aids future planning of similar studies. While some people chose not to participate due to concerns about undergoing a muscle biopsy, it is reassuring to note that over half of participants who consented to this procedure found it to be better than they expected and, most would be willing to undergo it again. This most likely reflects the high quality of care provided by clinical colleagues responsible for delivery of this important component of the study assessment.

By tracking recruitment and reporting these figures we provide transparency. This also allows us to consider how the characteristics of our study participants compare with those of the overall population from the areas in which they were recruited. Selection bias is an important consideration that some previous studies which include collection of muscle biopsy samples have overlooked.^15^ The comparisons we have undertaken highlight some evidence of selection bias, as indicated by the high levels of socioeconomic advantage within the study population. This is a common feature of population-based studies aiming to recruit community-dwelling adults. However, there are similar distributions of behavioural risk factors, most notably overweight and obesity and LTC, to other adults living in the same region, considerable inter-individual variability in health and functional status and a relatively high prevalence of MLTC. This suggests that our aim of developing an inclusive study, by purposefully restricting the number of exclusion criteria and recruiting as heterogeneous a sample as possible has been achieved, which is a major strength of our study, setting it apart from others.

Our flexible and agile approach enabled us to deliver a larger study covering a wider age range than originally planned. This allowed us to increase our statistical power and improve our ability to explore meaningful variations in ageing profiles across the full adult life course. This was achieved despite the COVID-19 pandemic and changes to the research team over time. We were also able to update study assessments in response to scientific advancements, for example, to update our assessment of MLTC and incorporate novel neurophysiology measures and more detailed assessments of muscle mass in a feasibility study. The latter demonstrates the potential value of this resource as a platform to support future innovation.

As we embark on a new phase of MASS Lifecourse and invite participants to undertake follow-up assessments, including a second muscle biopsy, approximately five to six years after their first assessment, this unique resource will be further enhanced. In the meantime, we aim to interrogate biological and environmental drivers of inter-individual differences in muscle ageing across the adult life course; an opportunity that can only be harnessed because of a major investment by many people and organisations including our study participants, research team, collaborators and funders.

## Data Availability

All data produced in the present study are available upon reasonable request to the authors

## Acknowledgements

The authors acknowledge their late colleague Dr Richard Dodds who was Principal Investigator of MASS Lifecourse until his death in May 2022. Richard played an instrumental role in the design and delivery of this study and the valuable resource generated forms part of his legacy.

The authors thank GP practices in the North East and North Cumbria Clinical Research Network including Northumbria Primary Care for their support of study recruitment and colleagues who supported study delivery including Terry Aspray, Matthew Birkbeck, Gayle Cain, Karen Davies, Kieren Hollingsworth, Sian Robinson, staff in the Clinical Ageing Research Unit (CARU) within the NIHR Newcastle Clinical Research Facility and radiographers within the Centre for In Vivo Imaging at Newcastle University.

The authors thank all those who have generously given their time (and muscle) to participate in MASS Lifecourse.

## Author contributions

AAS conceived the idea for this study, secured funding for fieldwork delivery and had overall responsibility for the design and delivery of the study. RC and MDW had responsibility for key elements of study delivery. CH led the fieldwork delivery team, supported by DC. CH, JGB, KSl, EGL, KSu and MDW delivered study visits. JGB, EGL, CM, KSu and MDW undertook muscle biopsies. AG contributed to study design and had oversight of the processing of biological samples. HA had responsibility for patient and public involvement and engagement work. SJH oversaw data management and SJH and HS undertook data cleaning and quality checks and prepared the master dataset. HS undertook the analyses presented. RC and HS drafted the manuscript. All authors critically reviewed and edited the manuscript and approved the final version for submission.

## Data sharing statement

The MASS Lifecourse Study Team supports data sharing with bona fide researchers and industry partners. Data and sample access is managed via an established collaboration and data request process in adherence with data security standards and in line with ethical approvals and participant consent. Contact details for enquiries are provided on the MASS Lifecourse study web page.^42^

---

**Supplementary Material – Table 1.** Participant experience of the muscle biopsy, and overall feedback on the MASS Lifecourse baseline study by sex.

| **n (%)<sup>\*</sup>** | **Male** | **Female** |
| --- | --- | --- |
| ***Participant experience of the muscle biopsy*** | ***n=115*** | ***n=123*** |
| **Experience relative to expectations** |  |  |
| Better than expected | 73 (63.5) | 61 (49.6) |
| As expected | 40 (34.8) | 59 (48.0) |
| Worse than expected | 2 (1.7) | 3 (2.4) |
| **Willing to have another for research** |  |  |
| Yes | 111 (96.5) | 120 (97.6) |
| No | 3 (2.6) | 3 (2.4) |
| Probably | 1 (0.9) | 0 (0.0) |
| ***Overall participant feedback on the study*** | ***n=118*** | ***n=126*** |
| **Interested in contributing to development of future research** |  |  |
| Yes | 117 (99.2) | 125 (99.2) |
| No | 1 (0.8) | 1 (0.8) |
| **Permission to contact for participation in future research** |  |  |
| Yes | 117 (99.2) | 125 (99.2) |
| No | 1 (0.8) | 1 (0.8) |
| **Interested to hear about the findings of MASS Lifecourse** |  |  |
| Yes | 118 (100.0) | 126 (100.0) |

<sup>\*</sup>Summary statistics are frequencies and percentages of non-missing sample size. 119 men and 127 women attended a MASS Lifecourse clinic assessment. The muscle biopsy was attempted for 116 men and 123 women, and all but 1 man rated their experience of the biopsy. One male and one female clinic attendee did not provide overall feedback on the study.

